# Hemodynamic Responses to Cold Water Ingestion During Head-Up Tilt

**DOI:** 10.64898/2026.09.29.26364369

**Authors:** Satoshi Kubota, Katsuya Yamauchi, Mitsue Kubota, Yutaka Endo

## Abstract

**Purpose:** We examined whether drinking a small volume of cold water during orthostatic stress attenuates the fall in systolic blood pressure (SBP).

**Methods:** In a randomized crossover study, 19 healthy young adults underwent 15 min of 60° head-up tilt under three conditions on separate days: no water, 200 mL of body-temperature water (37 °C), and 200 mL of cold water (4 °C). Drinking began 5 min after tilting and lasted 1 min. Hemodynamics and tympanic temperature were analyzed using covariate-adjusted linear mixed models with within- and between-condition comparisons.

**Results:** During drinking, SBP increased from pre-drinking values by 9.54 mmHg (95% confidence interval [CI] 5.58 to 13.50) with body-temperature water and 14.70 mmHg (10.74 to 18.66) with cold water. After cold water ingestion, SBP remained higher by 5.95 mmHg (95% CI 1.99 to 9.91) over the first 4 min and 4.84 mmHg (0.88 to 8.80) over the subsequent 5 min. No clear post-drinking increases occurred with body-temperature water. SBP was higher with cold water than with both other conditions during the first 4 min after drinking, and higher than with body-temperature water during the subsequent 5 min. After cold water ingestion, stroke volume was higher and heart rate lower than before drinking, and tympanic temperature declined between the two post-drinking periods.

**Conclusion:** Drinking 200 mL of cold water after blood pressure had fallen attenuated the reduction in SBP during continued orthostatic stress in healthy young adults.

## 1. Introduction

During orthostatic stress, gravity shifts blood to the legs and abdomen, reducing venous return. In healthy individuals, baroreflex-mediated sympathetic activation increases heart rate and causes peripheral vasoconstriction, usually minimizing the fall in blood pressure. When these compensatory responses are inadequate, orthostatic hypotension or orthostatic intolerance can develop, causing dizziness, presyncope, or syncope (Palma and Kaufmann 2020; Kim and Farrell 2022).

Water drinking is a common nonpharmacological treatment for orthostatic hypotension (Palma and Kaufmann 2020; Oyewunmi et al. 2023). Head-up tilt (HUT) studies in healthy individuals and patients with orthostatic syncope have reported improved orthostatic tolerance after ingestion of 473–500 mL of water 5–15 min before tilting (Schroeder et al. 2002; Lu et al. 2003; Claydon et al. 2006). Rapid ingestion of approximately 500 mL of water is also recommended for neurogenic orthostatic hypotension (Palma and Kaufmann 2020). These interventions use the osmopressor response to water ingestion: blood pressure begins to rise within 5 min and peaks at 30–40 min (Jordan et al. 2000; May and Jordan 2011). However, drinking approximately 500 mL over a short period can be physically demanding for older adults and frail patients, limiting its feasibility (Kubota et al. 2022).

In addition to the osmopressor response, drinking can elicit an immediate pressor response through afferent input from mechanoreceptors in the muscles involved in swallowing (Abe et al. 2013; Endo et al. 2002). This response is thought to be transient and largely confined to drinking, but may be modified by water temperature. In a previous study in healthy individuals, ingestion of 200 mL of cold water produced a greater pressor response than room-temperature water, and the response persisted after drinking (Kubota et al. 2022). Cold stimulation may augment swallowing-related afferent input during drinking, whereas a reduction in core temperature may sustain the pressor response afterward. These findings suggest that even a small volume of water could help maintain blood pressure during orthostatic stress if its temperature and timing are appropriately chosen. A study of water temperature and orthostatic tolerance initiated tilting 15 min after drinking (Parsons et al. 2022), but used a large volume of 500 mL within the framework of the osmopressor response. Whether drinking a small volume during orthostatic stress can attenuate the fall in blood pressure, and whether this effect persists after drinking, remain unclear.

We compared three conditions in healthy young adults during HUT—no water, 200 mL of body-temperature water, and 200 mL of cold water—to determine whether drinking attenuates the fall in systolic blood pressure. We hypothesized that body-temperature water would produce a transient pressor response during drinking, whereas the response to cold water would persist during continued tilting after drinking.

## 2. Methods

### 2.1 Participants and study design

This randomized crossover trial included 19 healthy young adults with no history of cardiovascular or respiratory disease, no chronic illness affecting daily activities, and no regular medication use. There were 8 men and 11 women. Mean (standard deviation [SD]) age, height, body mass, and body mass index (BMI) were 20.84 (1.07) years, 163.02 (8.89) cm, 53.89 (10.28) kg, and 20.13 (2.20) kg/m^2^, respectively. Women were studied during the follicular phase because sympathetic activity, baroreflex sensitivity, and vascular responsiveness may vary across the menstrual cycle (Minson et al. 2000). The study followed the principles of the Declaration of Helsinki and was approved by the Research Ethics Committee of the International University of Health and Welfare (approval no. 24-TA-092). All participants received oral and written explanations of the study purpose and procedures and provided written informed consent. The trial was registered with the UMIN Clinical Trials Registry (UMIN000057980).

### 2.2 Experimental conditions and protocol

The experimental environment followed our previous studies (Kubota et al. 2015; Kubota et al. 2017). Measurements were performed in a quiet room maintained at 27–28 °C to minimize thermal stress. This range is consistent with the use of 28 °C as a thermoneutral control in human experiments (Nishimura et al. 2017). All sessions took place between 11:00 and 15:00 to minimize circadian effects. Participants were asked to avoid strenuous exercise, caffeine, and alcohol from the day before testing, to consume only water after dinner, and to refrain from all food and fluids for 2 h before the experiment. During measurements, they wore a sleeveless shirt and shorts over their underwear.

Fig. 1 shows the protocol and analysis periods. Participants completed three conditions on separate days in randomized order: no water, 200 mL of body-temperature water (approximately 37 °C), and 200 mL of cold water (approximately 4 °C). Participants were aware of the assigned drinking condition before arriving for each session and were not blinded. Body-temperature water was included as a control to minimize the direct thermal effect of ingested water on core temperature. Each session consisted of 10 min of supine rest (baseline), 15 min of passive 60° HUT, and a further 5 min of supine rest (recovery). In the two water conditions, drinking began 5 min after tilting. The investigator delivered water through a catheter tube connected to a syringe, and participants drank 200 mL over 1 min. For each participant and condition, hemodynamic and tympanic temperature values were averaged over each analysis period. The periods were defined as HUT1 (minutes 3–5 of tilting; 2 min), Drinking (minutes 5–6; 1 min), HUT2 (the 4 min immediately after Drinking), HUT3 (the subsequent 5 min), and Recovery (minutes 3–5 after return to the supine position; 2 min). In the no-water condition, hemodynamic observations from minutes 5–6 of tilting were included as the time-matched control for the Drinking period.

**Fig. 1.**
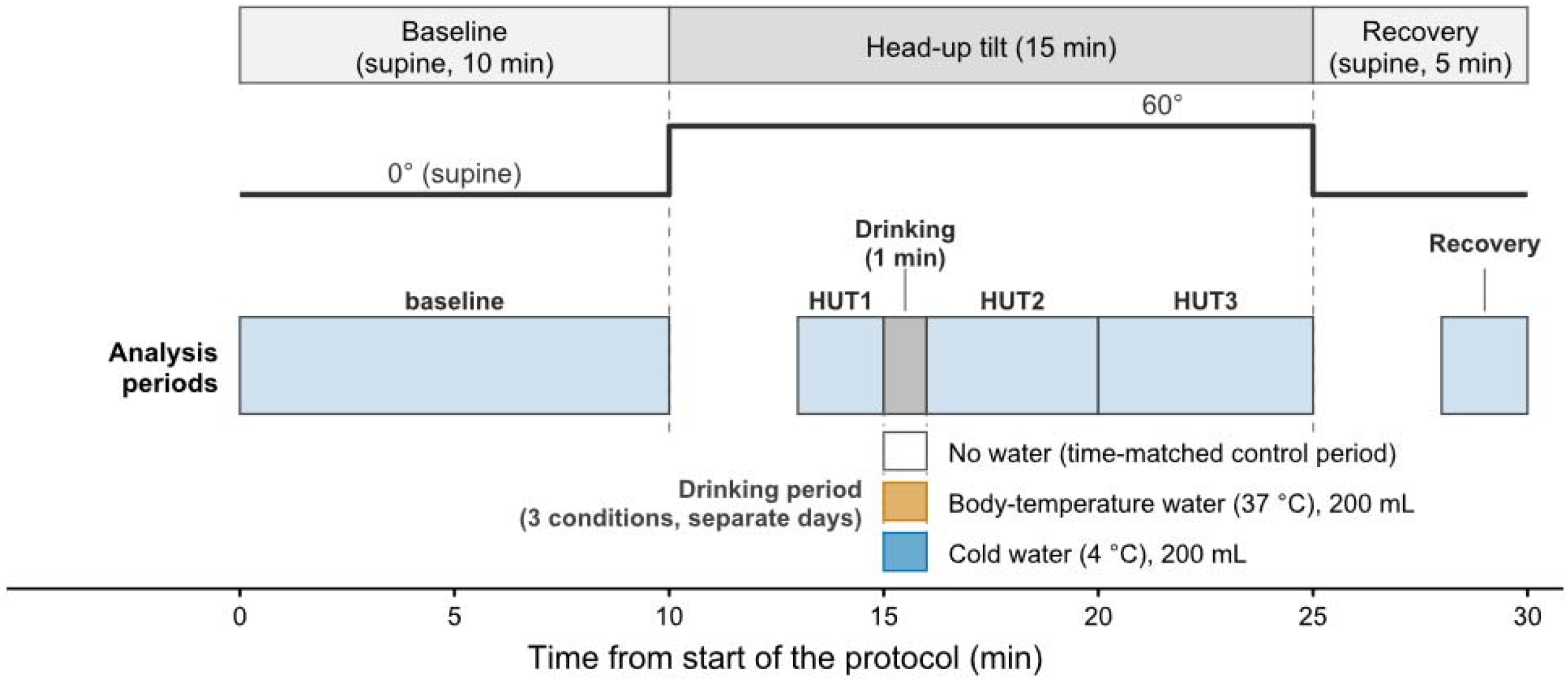
Experimental protocol and analysis periods. Participants underwent 10 min of supine rest, 15 min of 60° head-up tilt (HUT), and 5 min of supine recovery. Analysis periods were baseline (the entire 10-min supine rest), HUT1 (minutes 3–5 of tilt), Drinking (minutes 5–6), HUT2 (minutes 6–10), HUT3 (minutes 10–15), and Recovery (minutes 3–5 after returning to the supine position). During Drinking, participants consumed 200 mL of body-temperature water (37 °C) or cold water (4 °C) over 1 min. The corresponding interval in the no-water condition served as the time-matched control. The three conditions were performed on separate days in randomized order

### 2.3 Hemodynamic and tympanic temperature measurements

Beat-to-beat hemodynamics were recorded continuously using a noninvasive blood pressure monitor (Portapres, Finapres Medical Systems) with the cuff placed on the right index finger. The cuffed hand was kept at heart level throughout, including during HUT. Systolic blood pressure (SBP, mmHg), diastolic blood pressure (DBP, mmHg), mean blood pressure (MBP, mmHg), and heart rate (HR, beats/min) were derived from the arterial pressure waveform. Stroke volume (SV, mL) was estimated using the Modelflow method, and systemic vascular resistance (SVR, mmHg·s·mL□^1^) was calculated from the resulting hemodynamic measurements. Comparisons with Doppler ultrasound have shown systematic differences in Modelflow-derived SV during head-up tilt (van Lieshout et al. 2003). SV and SVR were used to assess relative changes rather than absolute values. SBP was the primary outcome; the other hemodynamic measures were secondary outcomes. Because the baroreflex maintains MBP during orthostasis in healthy individuals, we expected the effect of drinking to be most evident in SBP. Representative values for each predefined period were used in the analyses. Tympanic temperature was measured continuously as a noninvasive indicator of core temperature using a CE Thermo device (Nipro Corporation). Signals were acquired on a personal computer through a BIOPAC MP150 system at a sampling frequency of 1000 Hz.

### 2.4 Statistical analysis

Hemodynamic outcomes from HUT1, Drinking, HUT2, HUT3, and Recovery were analyzed using linear mixed models (LMMs) adjusted for baseline values. Fixed effects were condition, period, and their interaction. Random intercepts for participant and condition nested within participant accounted for repeated observations and differences in measurement levels between study days. Tympanic temperature was analyzed using the same model structure, with HUT1 immediately before drinking as the covariate and only HUT2 and HUT3 as outcomes to assess post-drinking temperature responses.

Observed reference values (baseline hemodynamics and HUT1 tympanic temperature) were summarized as means (SDs), excluding missing observations. Estimated marginal means (EMMs) and 95% confidence intervals (CIs) were evaluated at the covariate mean pooled across conditions and expressed as adjusted changes by subtracting that mean.

Hemodynamic analyses focused on within-condition contrasts of Drinking, HUT2, and HUT3 versus HUT1, with Dunnett-type adjustment within each outcome and condition. Recovery was excluded from these contrasts because posture differed from HUT1. For tympanic temperature, HUT3 was compared with HUT2 without adjustment because there was only one contrast per condition. For all outcomes, Tukey adjustment was applied to the three pairwise condition comparisons at each period. Adjustments applied to CIs and p values within each comparison family, with no additional correction across families or outcomes. Interpretation emphasized estimated differences and 95% CIs, with p values supplementary.

Kenward–Roger degrees of freedom were used for EMMs and contrasts. Analyses were performed in R 4.4.1 using lme4, lmerTest, pbkrtest, and emmeans. When an interrupted session was repeated, data from the repeat session were used for that condition.

### 2.5 Sample size

Sample size was estimated a priori for the primary outcome, SBP, using data from our previous study (Kubota et al. 2022). Because the pressor response during drinking had already been established, the calculation targeted persistence of the effect after drinking. An LMM was fitted to data from young participants in the cold-water condition of that study, with baseline SBP as a covariate, period as a fixed effect, and a participant-specific random intercept. The target was the increase from baseline at 5 min after drinking. Variance was inflated by a factor of 1.25 to allow for greater variability during HUT. With α set at 0.05 1000 Monte Carlo simulations were performed for each sample size to identify the size at which the lower limit of the 95% CI for power exceeded 80%. The required sample size was 17 participants. Allowing for potential dropout, the recruitment target was set at 19. Simulations used the simr package in R.

## 3. Results

### 3.1 Systolic blood pressure

All 19 enrolled participants were included in the analysis. In the no-water condition, two participants developed symptoms of acute orthostatic hypotension during HUT, and their sessions were stopped. In both participants, symptoms developed after the scheduled start of the Drinking period. One repeated the session on another day without these symptoms, and the repeat data were used. The other did not repeat the session, leaving no observations for the no-water condition. In another participant, equipment malfunction during the no-water session prevented data collection at HUT3 and Recovery. These interruptions and missing data occurred only in the no-water condition. Thus, the hemodynamic analyses included 18 participants at HUT1, Drinking, and HUT2 and 17 at HUT3 and Recovery in the no-water condition, and all 19 participants at every period in both water conditions. The results below are model-based estimates and 95% CIs adjusted for baseline values for hemodynamic outcomes and HUT1 values for tympanic temperature.

Table 1 presents observed baseline hemodynamic values and HUT1 tympanic temperatures for each condition. Figs. 2 and 3 show baseline-adjusted changes in SBP and the other hemodynamic measures, respectively.

**Table 1.** Observed baseline hemodynamic values and HUT1 tympanic temperature by drinking condition.

| Outcome | No water | Body-temperature water | Cold water |
| --- | --- | --- | --- |
| SBP (baseline), mmHg | 102.39 (10.16), n=18 | 102.53 (13.64), n=19 | 102.04 (11.66), n=19 |
| DBP (baseline), mmHg | 54.47 (8.00), n=18 | 54.79 (7.88), n=19 | 52.33 (9.19), n=19 |
| MBP (baseline), mmHg | 73.53 (9.22), n=18 | 73.48 (9.22), n=19 | 71.47 (9.82), n=19 |
| HR (baseline), beats/min | 62.35 (6.77), n=18 | 62.33 (6.08), n=19 | 61.72 (9.49), n=19 |
| SV (baseline), mL | 75.26 (13.53), n=18 | 75.92 (14.35), n=19 | 78.17 (11.94), n=19 |
| SVR (baseline), mmHg·s·mL<sup>-1</sup> | 0.97 (0.18), n=18 | 0.96 (0.15), n=19 | 0.93 (0.21), n=19 |
| Tympanic temperature (HUT1), °C | 36.87 (0.37), n=17 | 36.85 (0.24), n=19 | 36.86 (0.31), n=19 |

Note: Values are observed means (SD); n denotes the number of non-missing observations. Hemodynamic values were obtained at baseline and tympanic temperature at HUT1. No between-condition hypothesis tests were performed for these values. EMMs were evaluated at the pooled mean across all three conditions for the corresponding baseline or HUT1 covariate. SBP, systolic blood pressure; DBP, diastolic blood pressure; MBP, mean blood pressure; HR, heart rate; SV, stroke volume; SVR, systemic vascular resistance; HUT, head-up tilt; CI, confidence interval; EMM, estimated marginal mean; SD, standard deviation.

**Fig. 2.**
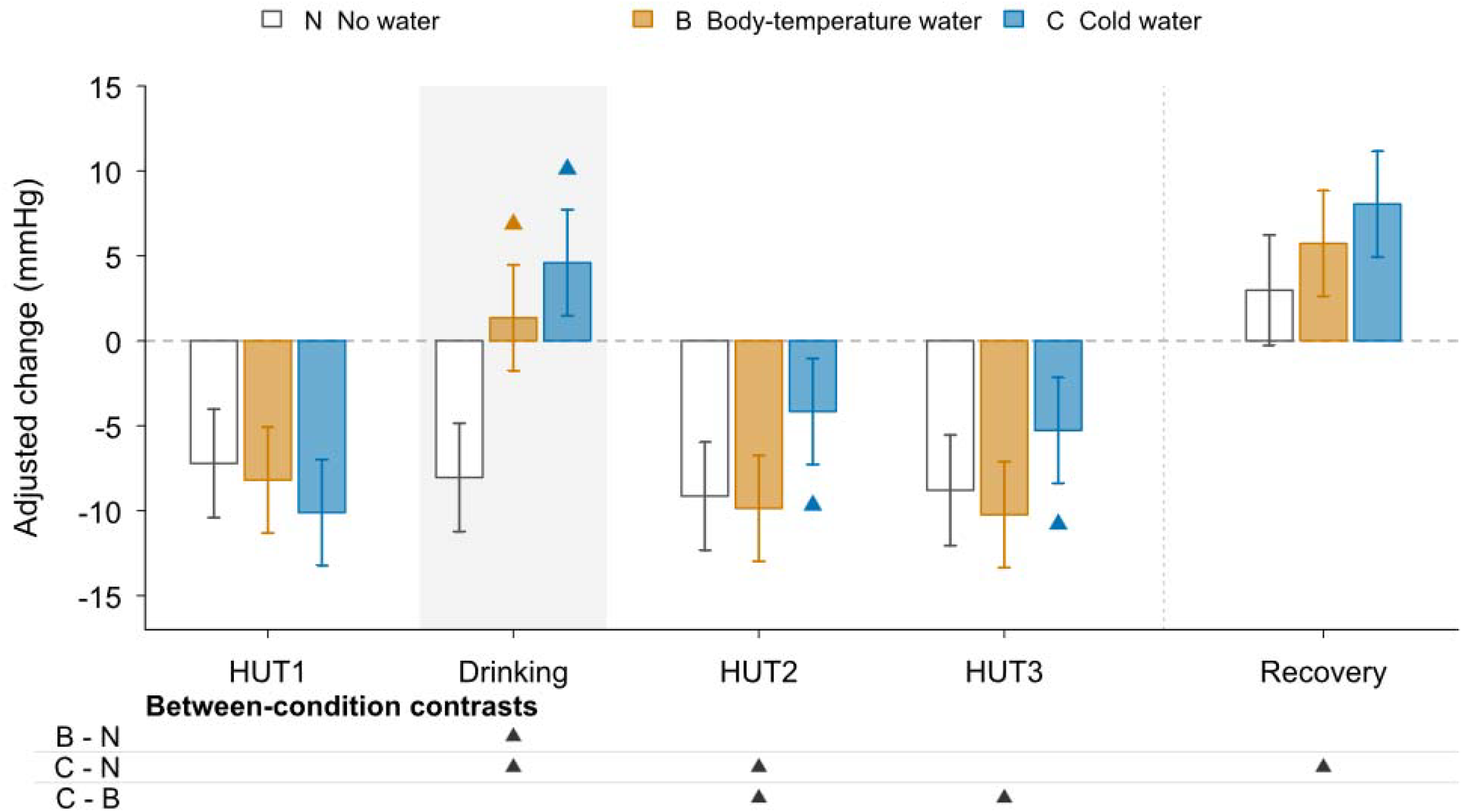
Baseline-adjusted changes in systolic blood pressure during head-up tilt and recovery. Bars show estimated marginal means evaluated at the pooled baseline covariate mean and expressed as changes from that mean; error bars indicate 95% confidence intervals (CIs). Triangles mark contrasts whose multiplicity-adjusted 95% CIs exclude zero: Dunnett adjustment for within-condition comparisons versus HUT1 and Tukey adjustment for between-condition comparisons. Within-condition contrasts use HUT1 as reference and exclude Recovery. Grey band: Drinking (time-matched for N). Available n: N=18 at HUT1/Drinking/HUT2 and 17 thereafter; B/C=19 throughout. Contrast estimates and CIs: Table 2. N, no water; B, body-temperature water; C, cold water. Coloured triangles indicate within-condition comparisons; black triangles indicate between-condition comparisons. Upward and downward triangles indicate positive and negative contrasts, respectively

**Fig. 3.**
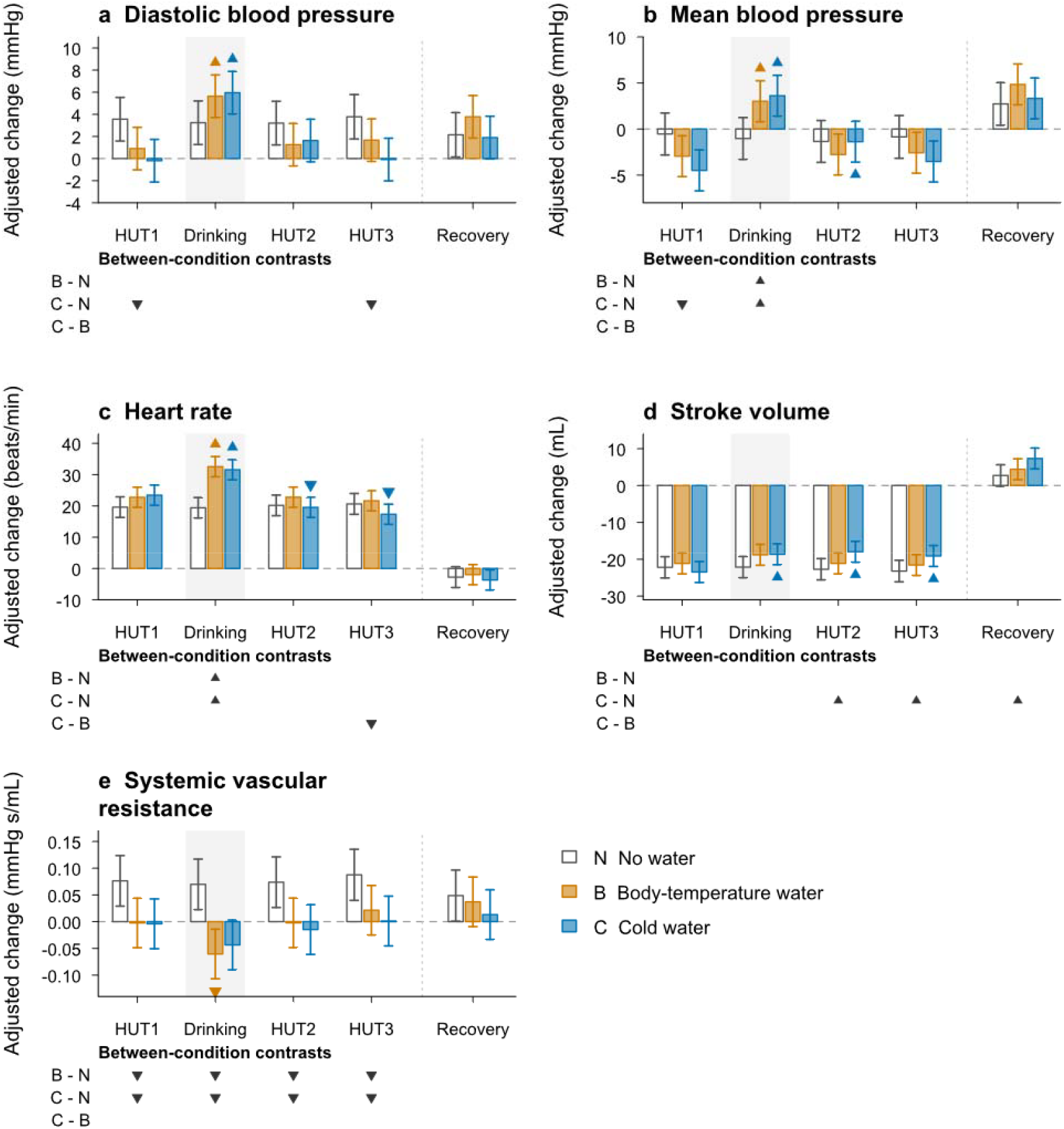
Baseline-adjusted changes in other hemodynamic measures during head-up tilt and recovery. Bars show estimated marginal means evaluated at the pooled baseline covariate mean and expressed as changes from that mean; error bars indicate 95% confidence intervals (CIs). Coloured triangles indicate within-condition comparisons using Dunnett adjustment; black triangles indicate between-condition comparisons using Tukey adjustment. Upward and downward triangles indicate positive and negative contrasts, respectively. Within-condition contrasts use HUT1 as reference and exclude Recovery. Grey band: Drinking (time-matched for N). Available n: N=18 at HUT1/Drinking/HUT2 and 17 thereafter; B/C=19 throughout. Contrast estimates and CIs: Table 3. N, no water; B, body-temperature water; C, cold water. HUT, head-up tilt. Panels: a, diastolic blood pressure; b, mean blood pressure; c, heart rate; d, stroke volume; e, systemic vascular resistance

Tables 2 and 3 report estimated differences, multiplicity-adjusted 95% CIs, and adjusted p values for within-condition contrasts versus HUT1 and between-condition contrasts at each period.

**Table 2.** Primary outcome (systolic blood pressure, mmHg): within-condition changes from HUT1 and between-condition comparisons.

| Period | Within-condition change from HUT1 (95% CI), p value |  |  |
| --- | --- | --- | --- |
|  | No water | Body-temperature water | Cold water |
| **HUT1** | Reference | Reference | Reference |
| **Drinking** | -0.83 (-4.90 to 3.24), p = 0.902 | 9.54 (5.58 to 13.50), p < 0.001 | 14.70 (10.74 to 18.66), p < 0.001 |
| **HUT2** | -1.92 (-5.99 to 2.14), p = 0.533 | -1.67 (-5.63 to 2.29), p = 0.612 | 5.95 (1.99 to 9.91), p = 0.001 |
| **HUT3** | -1.58 (-5.72 to 2.56), p = 0.672 | -2.04 (-6.00 to 1.92), p = 0.470 | 4.84 (0.88 to 8.80), p = 0.012 |
| **Recovery** | — | — | — |
| Period | Between-condition contrast, estimated difference (95% CI), p value |  |  |
|  | Body-temperature water - No water | Cold water - No water | Cold water - Body-temperature water |
| **HUT1** | -0.98 (-5.64 to 3.69), p = 0.873 | -2.89 (-7.56 to 1.77), p = 0.309 | -1.92(-6.51 to 2.68), p = 0.586 |
| **Drinking** | 9.39 (4.73 to 14.06), p < 0.001 | 12.64 (7.97 to 17.30), p < 0.001 | 3.24(-1.35 to 7.84), p = 0.220 |
| **HUT2** | -0.72 (-5.38 to 3.94), p = 0.929 | 4.98 (0.32 to 9.64), p = 0.033 | 5.70 (1.11 to 10.29), p = 0.011 |
| **HUT3** | -1.44 (-6.16 to 3.29), p = 0.753 | 3.52 (-1.20 to 8.25), p = 0.185 | 4.96 (0.37 to 9.55), p = 0.031 |
| **Recovery** | 2.75 (-1.97 to 7.48), p = 0.354 | 5.07 (0.34 to 9.80), p = 0.032 | 2.32 (-2.28 to 6.91), p = 0.458 |

Note: SBP is in mmHg. Upper block: within-condition contrasts versus HUT1, with Dunnett-adjusted simultaneous 95% CIs and p values. Recovery was excluded because posture differed from HUT1. Lower block: between-condition contrasts at each period, with Tukey-adjusted 95% CIs and p values; positive differences indicate higher values in the first-listed condition. During Drinking, no water served as the time-matched control. Available n: no water, 18 at HUT1, Drinking, and HUT2, and 17 at HUT3 and Recovery; both water conditions, 19 throughout. Adjusted changes from baseline are shown in Fig. 2. CI, confidence interval; HUT, head-up tilt; SBP, systolic blood pressure.

**Table 3.** Secondary hemodynamic outcomes: within-condition comparisons with HUT1 Secondary hemodynamic outcomes: between-condition comparisons.

| Outcome | Period | Within-condition change from HUT1 (95% CI), p value |  |  |
| --- | --- | --- | --- | --- |
|  |  | No water | Body-temperature water | Cold water |
| **DBP** | HUT1 | Reference | Reference | Reference |
|  | Drinking | -0.32 (-2.87 to 2.24), p = 0.965 | 4.74 (2.25 to 7.23), p < 0.001 | 6.15 (3.66 to 8.64), p < 0.001 |
|  | HUT2 | -0.35 (-2.91 to 2.21), p = 0.957 | 0.35 (-2.13 to 2.84), p = 0.953 | 1.82 (-0.67 to 4.31), p = 0.205 |
|  | HUT3 | 0.22 (-2.38 to 2.82), p = 0.984 | 0.76 (-1.73 to 3.25), p = 0.782 | 0.10 (-2.38 to 2.59), p = 0.996 |
|  | Recovery | — | — | — |
| **MBP** | HUT1 | Reference | Reference | Reference |
|  | Drinking | -0.49 (-3.47 to 2.50), p = 0.938 | 5.96 (3.06 to 8.86), p < 0.001 | 8.10 (5.20 to 11.00), p < 0.001 |
|  | HUT2 | -0.80 (-3.79 to 2.18), p = 0.828 | 0.18 (-2.72 to 3.08), p = 0.992 | 3.12 (0.22 to 6.02), p = 0.031 |
|  | HUT3 | -0.31 (-3.35 to 2.72), p = 0.976 | 0.37 (-2.53 to 3.27), p = 0.962 | 0.97 (-1.94 to 3.87), p = 0.744 |
|  | Recovery | — | — | — |
| **HR** | HUT1 | Reference | Reference | Reference |
|  | Drinking | -0.21 (-4.00 to 3.59), p = 0.993 | 9.78 (6.09 to 13.48), p < 0.001 | 8.12 (4.43 to 11.82), p < 0.001 |
|  | HUT2 | 0.56 (-3.23 to 4.36), p = 0.949 | 0.04 (-3.66 to 3.73), p = 1.000 | -3.88 (-7.57 to -0.19), p = 0.037 |
|  | HUT3 | 1.03 (-2.83 to 4.89), p = 0.832 | -1.10 (-4.80 to 2.59), p = 0.791 | -6.10 (-9.79 to -2.40), p < 0.001 |
|  | Recovery | — | — | — |
| **SV** | HUT1 | Reference | Reference | Reference |
|  | Drinking | 0.06 (-3.07 to 3.19), p = 0.999 | 2.34 (-0.70 to 5.38), p = 0.173 | 4.82 (1.78 to 7.86), p < 0.001 |
|  | HUT2 | -0.52 (-3.65 to 2.60), p = 0.934 | 0.00 (-3.04 to 3.05), p = 1.000 | 5.49 (2.45 to 8.54), p < 0.001 |
|  | HUT3 | -1.02 (-4.20 to 2.17), p = 0.763 | -0.44 (-3.48 to 2.61), p = 0.953 | 4.35 (1.30 to 7.39), p = 0.002 |
|  | Recovery | — | — | — |
| **SVR** | HUT1 | Reference | Reference | Reference |
|  | Drinking | -0.007 (-0.052 to 0.039), p = 0.951 | -0.058 (-0.102 to -0.014), p = 0.006 | -0.039 (-0.084 to 0.005), p = 0.091 |
|  | HUT2 | -0.003 (-0.048 to 0.043), p = 0.993 | 0.000 (-0.044 to 0.044), p = 1.000 | -0.011 (-0.055 to 0.033), p = 0.861 |
|  | HUT3 | 0.011 (-0.035 to 0.057), p = 0.860 | 0.024 (-0.020 to 0.068), p = 0.437 | 0.005 (-0.039 to 0.049), p = 0.968 |
|  | Recovery | — | — | — |

**Table 3 (continued). Secondary hemodynamic outcomes: between-condition comparisons**
| Outcome | Period | Between-condition contrast, estimated difference (95% CI), p value |  |  |
| --- | --- | --- | --- | --- |
|  |  | Body-temperature water - No water | Cold water - No water | Cold water - Body-temperature water |
| **DBP** | HUT1 | -2.65 (-5.64 to 0.33), p = 0.092 | -3.75 (-6.75 to -0.75), p = 0.010 | -1.10 (-4.05 to 1.86), p = 0.655 |
|  | Drinking | 2.40 (-0.58 to 5.38), p = 0.141 | 2.71 (-0.28 to 5.71), p = 0.085 | 0.32 (-2.64 to 3.27), p = 0.965 |
|  | HUT2 | -1.95 (-4.93 to 1.03), p = 0.272 | -1.58 (-4.58 to 1.42), p = 0.428 | 0.37 (-2.59 to 3.33), p = 0.953 |
|  | HUT3 | -2.11 (-5.14 to 0.91), p = 0.226 | -3.87 (-6.90 to -0.83), p = 0.008 | -1.75 (-4.71 to 1.20), p = 0.342 |
|  | Recovery | 1.63 (-1.39 to 4.65), p = 0.411 | -0.26 (-3.29 to 2.78), p = 0.978 | -1.88 (-4.84 to 1.07), p = 0.290 |
| **MBP** | HUT1 | -2.40 (-5.82 to 1.01), p = 0.221 | -3.95 (-7.38 to -0.52), p = 0.019 | -1.54 (-4.92 to 1.83), p = 0.526 |
|  | Drinking | 4.04 (0.63 to 7.45), p = 0.016 | 4.64 (1.21 to 8.06), p = 0.005 | 0.60 (-2.78 to 3.97), p = 0.908 |
|  | HUT2 | -1.42 (-4.83 to 1.99), p = 0.587 | -0.03 (-3.45 to 3.40), p = 1.000 | 1.40 (-1.98 to 4.77), p = 0.591 |
|  | HUT3 | -1.72 (-5.18 to 1.74), p = 0.469 | -2.67 (-6.14 to 0.80), p = 0.166 | -0.95 (-4.32 to 2.42), p = 0.783 |
|  | Recovery | 2.11 (-1.34 to 5.57), p = 0.320 | 0.59 (-2.87 to 4.06), p = 0.913 | -1.52 (-4.89 to 1.85), p = 0.537 |
| **HR** | HUT1 | 3.14 (-1.04 to 7.33), p = 0.181 | 3.82 (-0.37 to 8.01), p = 0.082 | 0.68 (-3.45 to 4.80), p = 0.921 |
|  | Drinking | 13.14 (8.95 to 17.32), p < 0.001 | 12.15 (7.96 to 16.34), p < 0.001 | -0.98 (-5.11 to 3.14), p = 0.839 |
|  | HUT2 | 2.62 (-1.57 to 6.81), p = 0.303 | -0.62 (-4.81 to 3.57), p = 0.935 | -3.24 (-7.36 to 0.88), p = 0.154 |
|  | HUT3 | 1.01 (-3.24 to 5.25), p = 0.841 | -3.31 (-7.56 to 0.95), p = 0.160 | -4.32 (-8.44 to -0.19), p = 0.038 |
|  | Recovery | 0.80 (-3.45 to 5.04), p = 0.898 | -0.91 (-5.17 to 3.34), p = 0.868 | -1.71 (-5.83 to 2.42), p = 0.591 |
| **SV** | HUT1 | 1.05 (-2.76 to 4.87), p = 0.790 | -1.27 (-5.10 to 2.55), p = 0.710 | -2.33 (-6.10 to 1.44), p = 0.312 |
|  | Drinking | 3.33 (-0.48 to 7.14), p = 0.100 | 3.49 (-0.34 to 7.32), p = 0.082 | 0.15 (-3.61 to 3.92), p = 0.995 |
|  | HUT2 | 1.58 (-2.23 to 5.39), p = 0.589 | 4.74 (0.91 to 8.57), p = 0.011 | 3.16 (-0.61 to 6.93), p = 0.119 |
|  | HUT3 | 1.63 (-2.23 to 5.49), p = 0.576 | 4.09 (0.22 to 7.96), p = 0.036 | 2.46 (-1.31 to 6.22), p = 0.274 |
|  | Recovery | 1.69 (-2.17 to 5.55), p = 0.553 | 4.61 (0.74 to 8.49), p = 0.015 | 2.92 (-0.85 to 6.69), p = 0.161 |
| **SVR** | HUT1 | -0.079 (-0.141 to -0.016), p = 0.010 | -0.080 (-0.143 to -0.017), p = 0.008 | -0.002 (-0.063 to 0.060), p = 0.998 |
|  | Drinking | -0.130 (-0.193 to -0.068), p < 0.001 | -0.113 (-0.176 to -0.050), p < 0.001 | 0.017 (-0.045 to 0.079), p = 0.792 |
|  | HUT2 | -0.076 (-0.139 to -0.014), p = 0.013 | -0.089 (-0.151 to -0.026), p = 0.003 | -0.013 (-0.074 to 0.049), p = 0.879 |
|  | HUT3 | -0.066 (-0.129 to -0.003), p = 0.037 | -0.086 (-0.150 to -0.023), p = 0.005 | -0.020 (-0.082 to 0.042), p = 0.718 |
|  | Recovery | -0.012 (-0.075 to 0.052), p = 0.900 | -0.035 (-0.099 to 0.028), p = 0.380 | -0.024 (-0.086 to 0.038), p = 0.628 |

Note: Units: DBP/MBP, mmHg; HR, beats/min; SV, mL; SVR, mmHg·s·mL<sup>-1</sup>. Within-condition contrasts: Dunnett-adjusted simultaneous 95% CIs/p values; Recovery excluded (different posture). Between-condition contrasts: Tukey-adjusted 95% CIs/p values; first-listed minus second-listed condition. No water is the time-matched Drinking control. Available n: no water, 18 (HUT1/Drinking/HUT2), 17 (HUT3/Recovery); both water conditions, 19 throughout. EMMs: Fig. 3. SBP, systolic blood pressure; DBP, diastolic blood pressure; MBP, mean blood pressure; HR, heart rate; SV, stroke volume; SVR, systemic vascular resistance; HUT, head-up tilt; CI, confidence interval; EMM, estimated marginal mean.

At HUT1, adjusted changes in SBP from baseline were −7.22 mmHg (95% CI −10.41 to −4.02) with no water, −8.19 mmHg (−11.31 to −5.07) with body-temperature water, and −10.11 mmHg (−13.23 to −6.99) with cold water, indicating a fall in SBP during HUT in all conditions (Fig. 2).

Within the no-water condition, there were no clear differences from HUT1 at Drinking, HUT2, or HUT3. With body-temperature water, SBP was higher during Drinking than at HUT1 (difference 9.54 mmHg, 95% CI 5.58 to 13.50), but the differences at HUT2 (difference −1.67 mmHg, −5.63 to 2.29) and HUT3 (difference −2.04 mmHg, −6.00 to 1.92) were not clear. With cold water, SBP was higher than at HUT1 during Drinking (difference 14.70 mmHg, 10.74 to 18.66) and remained higher at HUT2 (difference 5.95 mmHg, 1.99 to 9.91) and HUT3 (difference 4.84 mmHg, 0.88 to 8.80) (Table 2).

Between-condition comparisons showed higher SBP during Drinking with both body-temperature water (difference 9.39 mmHg, 95% CI 4.73 to 14.06) and cold water (difference 12.64 mmHg, 7.97 to 17.30) than with no water, with no clear difference between the two water temperatures. At HUT2, SBP was higher with cold water than with no water (difference 4.98 mmHg, 0.32 to 9.64) or body-temperature water (difference 5.70 mmHg, 1.11 to 10.29). At HUT3, SBP was higher with cold water than with body-temperature water (difference 4.96 mmHg, 0.37 to 9.55). At Recovery, SBP was higher with cold water than with no water (difference 5.07 mmHg, 0.34 to 9.80). No other between-condition differences were clear (Table 2).

### 3.2 Other hemodynamic measures

Baseline-adjusted changes in the other hemodynamic measures are shown in Fig. 3. At HUT1, adjusted HR estimates were above the pooled baseline reference, whereas adjusted SV estimates were below it in all three conditions.

Within-condition comparisons showed that DBP, MBP, and HR were higher during Drinking than at HUT1 in both water conditions, whereas SVR was lower with body-temperature water. With cold water, SV was also higher during Drinking than at HUT1 (difference 4.82 mL, 95% CI 1.78 to 7.86). The subsequent responses differed by water temperature. With body-temperature water, none of these outcomes clearly differed from HUT1 at HUT2 or HUT3. With cold water, SV was higher at HUT2 (difference 5.49 mL, 95% CI 2.45 to 8.54) and HUT3 (difference 4.35 mL, 1.30 to 7.39), whereas HR was lower at HUT2 (difference −3.88 beats/min, −7.57 to −0.19) and HUT3 (difference −6.10 beats/min, −9.79 to −2.40). MBP was also higher at HUT2 (difference 3.12 mmHg, 0.22 to 6.02). In the no-water condition, no outcome clearly differed from HUT1 at any of these periods. After drinking, SVR did not clearly differ from HUT1 in any condition (Table 3).

Between-condition comparisons showed higher HR and MBP and lower SVR during Drinking in both water conditions than in the no-water condition. SV was higher with cold water than with no water at HUT2 (difference 4.74 mL, 95% CI 0.91 to 8.57), HUT3 (difference 4.09 mL, 0.22 to 7.96), and Recovery (difference 4.61 mL, 0.74 to 8.49). At HUT3, HR was lower with cold water than with body-temperature water (difference −4.32 beats/min, 95% CI −8.44 to −0.19). SV did not clearly differ between the two water temperatures at any period (Table 3). Before drinking, at HUT1, DBP was higher with no water than with cold water, and SVR was higher with no water than in either water condition (Table 3).

### 3.3 Tympanic temperature

The tympanic temperature analysis included 17 participants in the no-water condition: one had missing data because of the session termination for orthostatic hypotension described above, and another because the temperature sensor became dislodged. Both water conditions included all 19 participants.

Adjusted changes in tympanic temperature from HUT1 are shown on the left side of Table 4, Panel A, and within-condition differences between HUT3 and HUT2 on the right. Panel B presents between-condition differences at each period. At HUT2, estimates indicated increases with no water and body-temperature water and little change with cold water. At HUT3, estimates indicated increases with no water and body-temperature water and a decrease with cold water.

**Table 4.** Tympanic temperature: HUT1-adjusted estimated marginal means, within-condition comparison, and between-condition comparisons during head-up tilt Panel A. Estimated marginal means and within-condition comparison.

| Period | Adjusted changes from HUT1, estimated marginal means (95% CI) |  |  | Within-condition comparison with HUT2, estimated difference (95% CI), p value |  |  |
| --- | --- | --- | --- | --- | --- | --- |
|  | No water | Body-temperature water | Cold water | No water | Body-temperature water | Cold water |
| HUT 2 | 0.023 (0.003 to 0.043) | 0.033 (0.013 to 0.052) | 0.010 (-0.009 to 0.030) | Reference | Reference | Reference |
| HUT 3 | 0.050 (0.030 to 0.071) | 0.063 (0.044 to 0.083) | -0.062 (-0.082 to -0.043) | 0.027 (0.012 to 0.043), p = 0.001 | 0.031 (0.016 to 0.046), p < 0.001 | -0.073 (-0.088 to -0.058), p < 0.001 |

**Panel B. Between-condition comparisons at each period**
| Period | Between-condition comparison, estimated difference (95% CI), p value |  |  |
| --- | --- | --- | --- |
|  | Body-temperature water - No water | Cold water - No water | Cold water - Body-temperature water |
| HUT2 | 0.010 (-0.024 to 0.043), p = 0.762 | -0.013 (-0.046 to 0.021), p = 0.629 | -0.022 (-0.055 to 0.010), p = 0.224 |
| HUT3 | 0.013 (-0.020 to 0.046), p = 0.622 | -0.113 (-0.146 to -0.080), p < 0.001 | -0.126 (-0.158 to -0.093), p < 0.001 |

Note: Unit: tympanic temperature, °C. Panel A, left: EMMs evaluated at the pooled HUT1 covariate mean and expressed relative to that mean (95% CIs). These values are descriptive; no direct comparisons with observed HUT1 values were performed. Panel A, right: within-condition contrasts of HUT3 versus HUT2, with 95% CIs and p values. No multiplicity adjustment was applied because there was one contrast per condition. Panel B: between-condition contrasts at each period, with Tukey-adjusted 95% CIs and p values. Positive differences indicate higher values in the first-listed condition. Available n: no water, 17; both water conditions, 19 each. CI, confidence interval; EMM, estimated marginal mean; HUT, head-up tilt.

Within-condition comparisons showed higher tympanic temperature at HUT3 than at HUT2 with no water (difference 0.027 °C, 95% CI 0.012 to 0.043) and body-temperature water (difference 0.031 °C, 0.016 to 0.046), but lower temperature with cold water (difference −0.073 °C, −0.088 to −0.058).

There were no clear between-condition differences at HUT2. At HUT3, tympanic temperature was lower with cold water than with no water (difference −0.113 °C, 95% CI −0.146 to −0.080) or body-temperature water (difference −0.126 °C, −0.158 to −0.093).

## 4. Discussion

Drinking 200 mL of water during HUT increased SBP above HUT1 levels during drinking at both water temperatures. With cold water, SBP remained above HUT1 levels at HUT2 and HUT3 after drinking. Between-condition comparisons showed higher SBP with cold water than with no water or body-temperature water at HUT2, and higher SBP than with body-temperature water at HUT3. These findings indicate that cold water ingestion can attenuate the fall in SBP during orthostatic stress.

During drinking, SBP was higher than immediately before drinking at HUT1 in both water conditions, despite the upright posture that predisposes SBP to fall. The no-water condition showed no clear difference from HUT1 over the same period. HR was also higher in both water conditions, and SV was higher with cold water, suggesting that this transient pressor response arose from maintenance or augmentation of cardiac output. SVR did not increase and was lower than at HUT1 with body-temperature water, making generalized peripheral vasoconstriction alone an unlikely explanation. Because SVR is calculated from blood pressure and cardiac output, however, its lower value could itself result from increased cardiac output. In baroreceptor-denervated rats, Abe et al. (2013) demonstrated a role for afferent input from mechanoreceptors in muscles involved in drinking and for α1-adrenergic receptor-mediated vasoconstriction. Whether the same mechanism operated in our healthy participants with intact baroreflexes is unclear. The pressor response was numerically greater with cold water: SBP rose by approximately 15 mmHg from HUT1, compared with approximately 10 mmHg with body-temperature water. This pattern suggests that cold stimulation may have augmented the afferent input or cardiac sympathetic response, consistent with the greater pressor responses to swallowing cold water and cold carbonated water reported previously (Kubota et al. 2022).

Responses during continued tilting after drinking differed between body-temperature and cold water. With body-temperature water, neither SBP nor any other hemodynamic outcome clearly differed from HUT1 at HUT2 or HUT3, indicating that the response during drinking was transient. With cold water, SBP remained higher than at HUT1 throughout HUT2 and HUT3. In healthy individuals, the baroreflex increases HR to compensate for the orthostatic reduction in SV and maintain blood pressure. After cold water ingestion, SV was higher than at HUT1, whereas HR was lower. Neither outcome clearly differed from HUT1 with no water or body-temperature water. The recovery of SV and reduction in HR, together with sustained higher SBP, suggest improved circulatory stability during continued orthostatic stress after cold water ingestion. Although the effect of drinking in these healthy participants was most evident in SBP, MBP was also higher than at HUT1 immediately after cold water ingestion. These measures alone cannot establish the mechanism of the pressor response.

At HUT1, DBP was higher with no water than with cold water, and SVR was higher with no water than in either water condition (Table 3). Within-condition comparisons, however, showed a common pattern in the two water conditions: DBP was higher during drinking, with lower SVR in the body-temperature water condition, and differences from HUT1 were no longer clear after drinking. This pattern did not correspond to the between-condition differences. We therefore consider these differences to reflect systematic differences between study days rather than effects of drinking. Participants knew the day’s condition before arriving, so their anticipatory response to tilting may have differed in the no-water condition, which involved no intervention (Gregg et al. 1999). However, the no-water condition showed no increase in HR consistent with emotional sympathetic arousal. The combination of higher DBP and SVR with relatively lower HR may instead reflect day-to-day variation in vasomotor tone. Psychological stress was not assessed, so this explanation remains speculative.

The sustained attenuation of the fall in blood pressure agrees with our previous finding that blood pressure returned rapidly to baseline after room-temperature water, whereas the pressor response to cold water and cold carbonated water persisted into recovery (Kubota et al. 2022). That study proposed a mechanism involving a reduction in core temperature after cold water ingestion, drawing on Frank et al. (1997), who showed that lowering core temperature without cooling the skin increased both plasma norepinephrine and blood pressure. Although we did not measure rectal or esophageal temperature, we recorded tympanic temperature as a noninvasive indicator of core temperature. Tympanic temperature does not necessarily correspond exactly to rectal or esophageal temperature and can be influenced by local thermal conditions (Desruelle and Candas 2000; Del Coso et al. 2008). It should therefore be interpreted here as an index of temperature dynamics near the central nervous system rather than a direct measure of whole-body core temperature. At HUT2, adjusted estimates indicated increases with no water and body-temperature water and values close to HUT1 with cold water, although the between-condition differences were not clear. This pattern suggests that cold water may have begun to suppress the rise in tympanic temperature during HUT by HUT2. Within-condition comparisons showed higher temperature at HUT3 than at HUT2 with no water and body-temperature water, whereas only cold water showed a reduction (difference −0.073 °C, 95% CI −0.088 to −0.058). The divergence in temperature responses between conditions was clear at HUT3. The core cooling studied by Frank et al. and the tympanic temperature changes observed here differ in both the measure used and the magnitude of temperature change. Our tympanic temperature findings do not establish a causal relationship with the pressor response, but provide supporting evidence that the influence of cold stimulation persisted after drinking.

With no water and body-temperature water, tympanic temperature was higher at HUT3 than at HUT2 (differences 0.027 °C, 95% CI 0.012 to 0.043, and 0.031 °C, 0.016 to 0.046, respectively), presumably reflecting a temperature response to continued orthostatic stress. Reductions in central blood volume and baroreceptor unloading during orthostatic stress can modify heat-loss responses, including cutaneous vasodilation and sweating (Mack et al. 1995; Ogawa et al. 2017; Johnson et al. 2014). In the absence of a cooling stimulus, the small rise in tympanic temperature may therefore have resulted from suppression of heat loss during HUT or changes in local circulation in the head and neck.

Parsons et al. (2022) examined the preventive effect of adding a temperature component to the osmopressor response before orthostatic stress. Ingestion of 500 mL of cold water 15 min before tilting did not prolong time to presyncope, but maintained higher SBP and SV during tilting than room-temperature water. This supports our finding that cold water can help maintain circulation during orthostatic stress. Parsons et al. also reported lower HR and cardiac output, identifying attenuation of orthostatic tachycardia as a potential clinical benefit. In our study, HR after cold water ingestion was also lower than immediately before drinking at HUT1, despite differences in water volume and timing. However, Parsons et al. also observed an increase in SVR. Differences in water volume, timing, and the form of orthostatic stress may account for the differing responses.

The responses observed here cannot be explained solely by the typical osmopressor response. The rise in blood pressure during the 1-min drinking period does not match the slower onset of the osmopressor response after drinking (May and Jordan 2011). Moreover, SBP was higher with cold water than with body-temperature water at both HUT2 and HUT3, even though water volume and osmolality were identical in the two water conditions. This difference points to a temperature-related effect rather than an osmotic stimulus. We consider the responses to reflect an immediate effect of swallowing modified by water temperature.

The clinical relevance of this study lies in the attenuation of the fall in SBP from the onset of drinking even when water was given after SBP had already fallen during HUT, rather than preventively before tilting. In particular, SBP with cold water remained approximately 5–6 mmHg higher at HUT2 and HUT3 than at HUT1, recovering about half of the approximately 10-mmHg orthostatic fall. Cold water ingestion may therefore offer a means of supporting blood pressure after an orthostatic fall has occurred, in addition to its use before standing. Even with body-temperature water, SBP during drinking was approximately 9 mmHg higher than with no water. Our previous study likewise found a transient pressor response to 200 mL of room-temperature water during seated rest (Kubota et al. 2022). The consistent immediate response to water without a cold stimulus across different postures and circulatory conditions suggests that drinking itself elicits a transient pressor response. Although cold water appears more favorable for a sustained effect, body-temperature water may provide short-term blood pressure support when cold stimulation is undesirable. This study provides proof of concept that drinking 200 mL of water, particularly cold water, can attenuate the fall in SBP during orthostatic stress. As this was a physiological study in healthy young adults, clinical efficacy, safety, and feasibility need to be tested in patients with orthostatic hypotension.

## Limitations

This study has several limitations. First, sample size was based on a simplified model using only the cold-water condition from our previous study and did not match the model used in the final analysis. Nevertheless, the a priori power calculation targeted a within-condition increase after drinking, the same type of effect as the main comparison in this study. It does not establish adequate power for between-condition comparisons or interaction effects. Multiplicity adjustment was limited to each comparison family, so findings spanning multiple comparisons require care in interpretation. Secondary outcomes were interpreted as exploratory.

Second, because the conditions were studied on separate days, between-condition comparisons include day-to-day variation, adding uncertainty to their estimates. DBP and SVR already differed between conditions before drinking at HUT1 (Table 3). In within-condition contrasts, the constant difference in measurement level between study days represented by the random intercept cancels out. Differences between days in the time course or response to orthostatic stress do not. Third, participants were healthy young adults, and symptoms were not assessed, limiting generalization to patients with orthostatic hypotension. Fourth, replacing or excluding no-water data after orthostatic hypotension may have favored better-tolerated sessions and biased between-condition comparisons. The direction and magnitude of this potential bias remain uncertain.

## Abbreviations

BMI: Body mass index
CI: Confidence interval
DBP: Diastolic blood pressure
EMM: Estimated marginal mean
HR: Heart rate
HUT: Head-up tilt
LMM: Linear mixed model
MBP: Mean blood pressure
SBP: Systolic blood pressure
SD: Standard deviation
SV: Stroke volume
SVR: Systemic vascular resistance

## Statements and Declarations

### Author contributions

SK, KY and YE conceived and designed the research and edited and revised the manuscript. SK, MK and KY analyzed data. SK and MK performed the measurements and organized the data. SK drafted the manuscript. All authors read and approved the final manuscript.

### Ethics approval

This study was performed in accordance with the Declaration of Helsinki and approved by the Research Ethics Committee of the International University of Health and Welfare (approval no. 24-TA-092).

### Consent to participate

Written informed consent was obtained from all participants.

### Data availability

The de-identified analysis dataset is available from the corresponding author upon reasonable request.

